# The importance of the human factor during the evolution of SARS-CoV-2 pandemic: the successful case of the Italian strategy

**DOI:** 10.1101/2020.10.22.20215277

**Authors:** Cristina Scarpazza, Gianluca Musumeci, Andrea S. Camperio Ciani

## Abstract

In Italy, 311,364 cases and 35,851 deaths of people who tested positive for SARS-CoV-2 were registered as of September 29th, 2020. To avoid the spreading of the virus, mathematical models predicting the course of infection’s spread^1^ become the basis to plan stringent countermeasures. We applied a published algorithm to real data up to September 27^th^, modeling two scenarios where predicted and real data were compared: a conservative scenario with a lockdown still ongoing and a scenario reflecting what actually happened in Italy, where the lockdown has been removed. Results revealed that the number of individuals in life-threatening condition is much lower than predicted, as well as the number of symptomatic individuals. Contrarily, the number of asymptomatic individuals is much higher than predicted. This suggest that human beings are not passive victims, but active fighters able to change the course of the infection creating adaptive strategies against the infection’s spread.

## Introduction

In December 2019, the first case of Coronavirus Disease 2019 (COVID-19) was identified in Wuhan (China), caused by the Severe Acute Respiratory Syndrome Coronavirus-2 (SARS-CoV-2). As COVID-19 rapidly spread throughout several nations, in January 2020 the World Health Organization (WHO) declared a state of sanitary emergency and, at the beginning of March 2020, declared the SARS-CoV-2 infection pandemic. Approximately 5% of patients with COVID-19, and 20% of those hospitalized, experienced symptoms necessitating intensive care. Among patients hospitalized in the intensive care unit (ICU), the case fatality is up to 40%^2^. As to September 29th, the WHO reported 33,603,488 total cases and 1,007,438 deaths worldwide.

Italy has been severely affected. A total of 311,364 confirmed cases (5,152 individuals in a million) and 35,851 deaths (592 individuals in a million) of people who tested positive for SARS-CoV-2 were registered as of September 29th, 2020.

Effective long-term control of transmission depended on understanding of the mechanisms of SARS-CoV-2 transmission, particularly on the contribution of asymptomatic, pre-symptomatic and symptomatic transmission^3^. A recent review^4^ suggests that asymptomatic persons (40%-45% of those infected) can transmit SARS-CoV-2 for a timeframe longer than 14 days, thus increasing the likelihood of unnoticed and pervasive transmission of the virus. To mitigate virus’s spreading and limit the medical, social, psychological and economic impact of the outbreak, different research groups estimated the transmission dynamics and the efficacy of potential control measures^1, 5^. In particular, one paper^1^, aiming to predict the future course of infection’s spread and mortality rate, proposed a mathematical model where different possible scenarios of implementation of countermeasures were simulated.

This algorithm is relevant as it discriminates between detected and undetected cases of infection, between symptomatic and asymptomatic individuals, and between different severity of illness^1^. The entire population was thus partitioned in Susceptible individuals (S); Infected (I: asymptomatic undetected); Diagnosed (D: asymptomatic detected); Ailing (A: Symptomatic undetected); Recognized (R: Symptomatic detected); Threatened (T: In life threatening condition); Healed (H); Extinct (E: dead). The term SIDARTHE was thus coined to refer to this model^1^.

The results from this model have been a precious help for the Italian government to guide the strength of the implemented countermeasures. Indeed, being Italy the first affected country after China, Italians were devoid of examples from other nations of effective strategies implementation to mirror. The Italian strategy to minimize the long term transmission of the virus has been organized in three phases: phase 1 (61 days: March 11th - May 4th) was characterized by a nationwide compulsory lockdown. During this phase, nasopharyngeal swabs were performed on symptomatic individuals only. Phase 2 (42 days: May 4th - June 15th) was characterized by a partial re-opening, as individuals were allowed to see relatives and to move for work purposes. During this phase, nasopharyngeal swabs were performed at population level, with the aim to identify and isolate asymptomatic individuals. Phase 3 (June 15th-still ongoing) is characterized by the possibility to go outdoors and meet other people. Social distancing and face masks wearing, as well as hands and surfaces disinfections, remain compulsory.

The Italian model is now considered paradigmatic as, despite the high number of nasopharyngeal swabs performed daily, the number of new infected is now low, contrary to what is happening in other European countries. To estimate the real impact of the Italian strategy, it is thus important to understand whether the real data on infected individuals and individuals in life-threatening conditions fit the data predicted by the original model^1^.

The original model, although extremely advanced, has however several limitations. First, it did not include the actual number of nasopharyngeal swabs performed, but it considered them as a constant (the double of the COVID-19 positive detected individuals, calculated as the sum of the discovery rate values of symptomatic and asymptomatic individuals). Second, it includes the data of the first 45 days only, thus the prediction could be unstable. Third and most important, SIDARTHE is a dynamic but regular model. As such, it did not take into consideration potential distortive effects, as the impact of new medical discoveries.

## Methods

For a detailed description of the methods please refer to Giordano et al. 2020^1^. The methods we applied are exactly the same of the ones applied in by the original model^1^.

In the current paper, we applied the SIDARTHE model^1^, whose algorithm is free available online, to real data on COVID-19, extending the empirical data of six months, with the aim to understand whether the actual data reflected the ones predicted by SIDARTHE.

As for the original publication, we inferred the model parameters based on the official data (source: Protezione Civile and Ministero della Salute) about the evolution of the epidemic in Italy. While the original publication included data from 20 February 2020 to 5 April 2020 (day 45), we extended the included actual data to 27 September 2020 (day 227). Furthermore, we included the actual daily number of nasopharyngeal swabs that, as illustrated in Figure 1A, has been much higher than estimated within the original model and, congruently, the number of individuals who tested positives has been much lower than the constant 50% considered within the model (Figure 1B).

**Figure 1.**
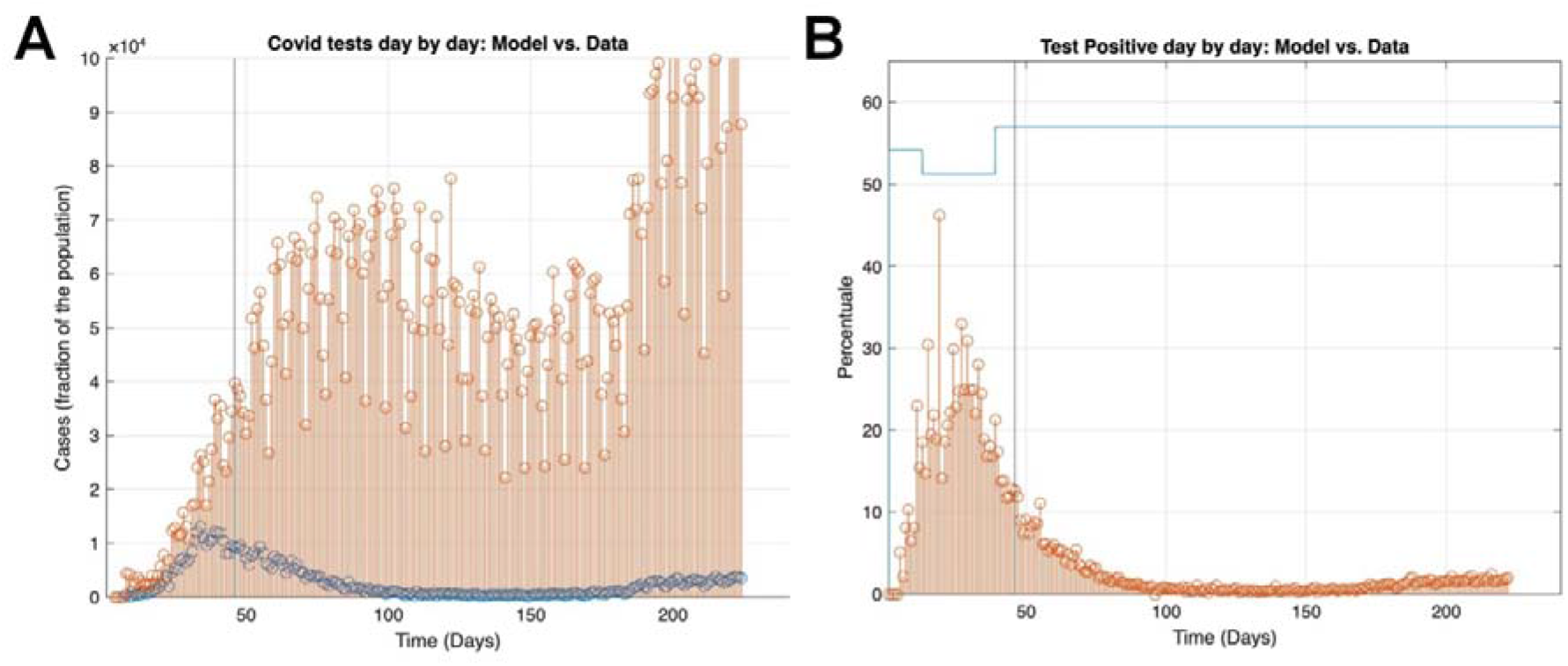
COVID-19 nasopharyngeal swabs. A: number of nasopharyngeal swabs every day. B: percentage of positive swabs per day. The vertical line on day 45 refers to the data included within the original publication^1^. In red: real data; In blue: data predicted by SIDARTHE.

Two different scenarios of implementation of countermeasures have been modeled in the current paper. First, we modeled an unrealistic scenario, where the lockdown is supposed to be still ongoing. To this end, we run the original SIDARTHE strengthened lockdown scenario algorithm, to test whether the predicted data using the most stringent countermeasures fit the real data till September 27th. Second, we created an alternative scenario where we reproduced what actually happened in Italy, where the lockdown was interrupted on May 5th, day 75 (phase 2). Hence, we modified the original SIDARTHE strengthened lockdown scenario algorithm to include the beginning of phase 2.

## Results and Discussion

The most important result is that, regardless of the scenario considered, the number of individuals in life threatening conditions (assisted in ICU) has been significantly lower than the one predicted by the original model (Figure 2A). Furthermore, the number of real deaths after day 80 has also been dramatically lower than the one predicted (Figure 2B). This latter data will not be extensively discussed as the authors of the original publication^1^ already highlighted that SIDARTHE prediction did not fit the data regarding deaths.

**Figure 2.**
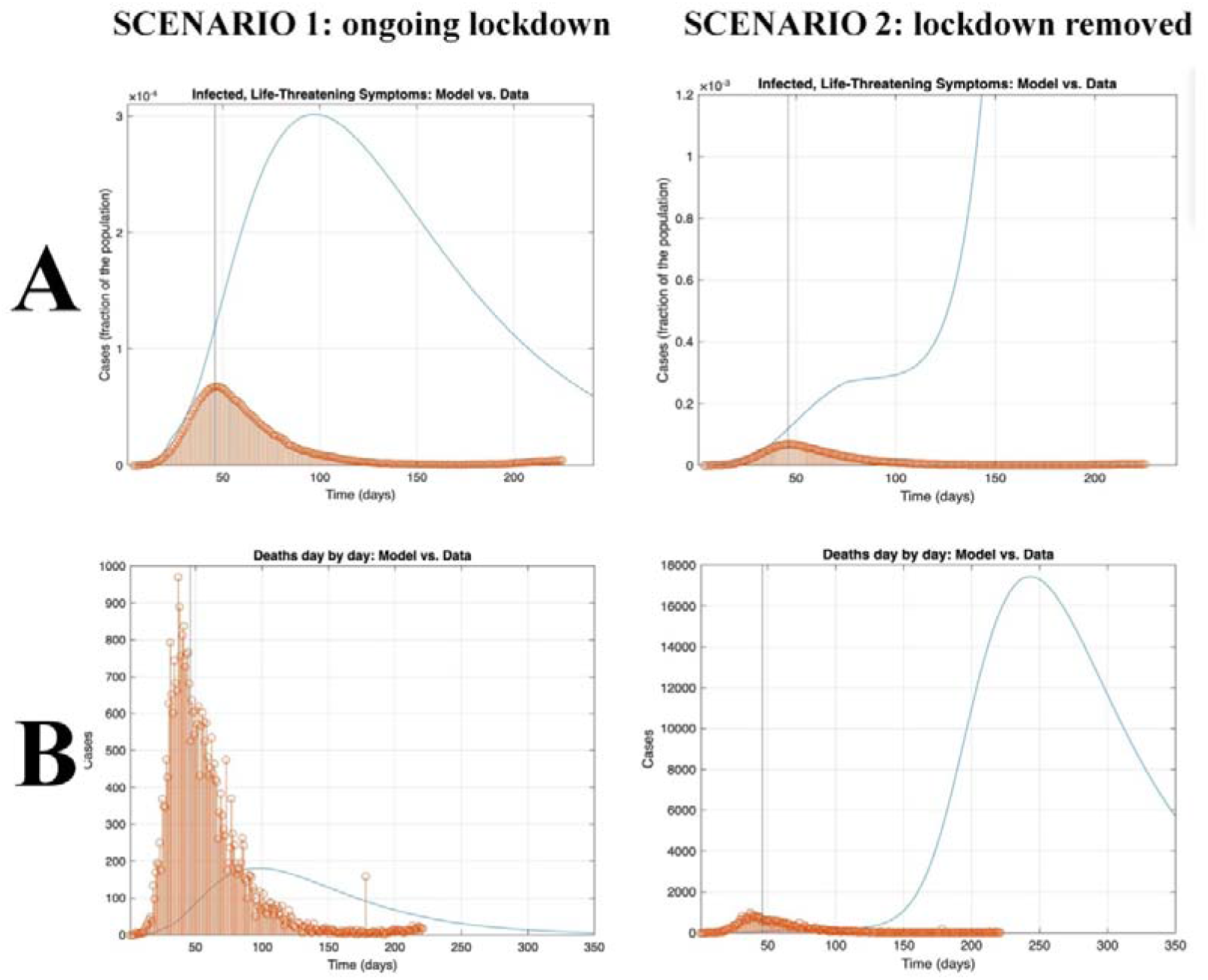
Effect of two countermeasures scenarios on individuals admitted to ICU and deaths. The figure represented the comparison between real data (red) and predicted data (blue) in the two scenarios: with ongoing lockdown (left) and removing lockdown (right). A) number of individuals in life threatening condition and admitted to ICU; B) number of deaths. The empirical data are exactly the same in the two different scenarios, while the y axis scale is different for illustrative purposes only.

These data suggest the important impact of emerging therapies to prevent excessive inflammation of the lungs, as the interleukin blockade, for instance anakinra (IL-1)^6^ or tocilizumab (IL-6). This latter has been found to reduce the mortality rate of 96% if administered early in the course of the disease, preventing excessive lung inflammation, and the consequent need to be admitted to ICU^7^. The identification of new therapeutic agents was coupled with the publication of evidence showing no benefit of initial standard therapies as cloroquine^8^, thus allowing clinicians to focus on effective therapies.

Importantly, just after the publication of the original SIDARTHE paper, the first autopsy series was published^9^, revealing an high incidence of thrombotic complication^10^. This insight was particularly relevant as it was followed by the administration of anticoagulants, resulting in a relevant decrease of admission to ICU, need of mechanical ventilation, and deaths^11^. Not less relevant, people are now paying more attention to COVID-19 symptoms, they are admitted to hospitals earlier during the disease course and, thus, can receive the right treatment earlier than before.

As second critical result, our data reveal that, regardless the scenario, the real number of asymptomatic COVID-19 positive individuals has been much higher than the one predicted by the original model, at least till day 100 (Figure 3A), while the real number of symptomatic COVID-19 positive individuals has been much lower than the one predicted (Figure 3B). Thus, the real data appeared to be the opposite of the one predicted by SIDARTHE.

**Figure 3.**
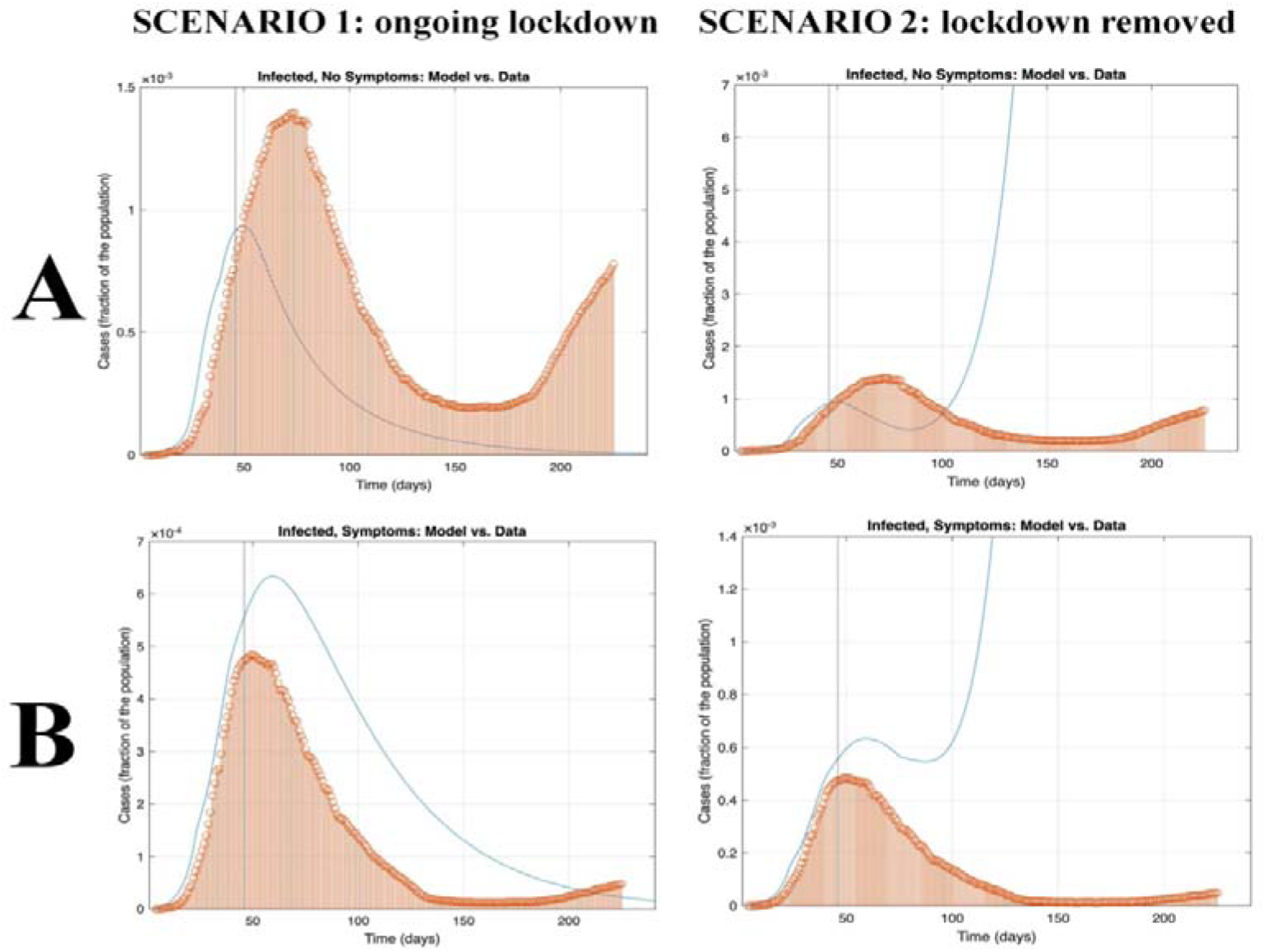
Effect of two countermeasures scenarios on asymptomatic and symptomatic COVID-19 individuals. The figure represented the comparison between real data (red) and predicted data (blue) in the two scenarios: with ongoing lockdown (left) and removing lockdown (right). A) number of asymptomatic individuals; B) number of symptomatic individuals. The empirical data are exactly the same in the two different scenarios, while the y axis scale is different for illustrative purposes only.

This result cannot be explained by medical improvement, as medical assistance begins with symptomatic individuals. The number of asymptomatic which increased much more than the number of symptomatic patients, coupled with a decrease of people in life-threatening condition, strongly suggests an evolutionary virus variants selection, that progressively evolves toward sub-lethal and asymptomatic expressions. This idea is supported by evidences showing two region of the virus that are under selective pressure, making the virus instable and prone to evolutionary changes^12^. Indeed, many viruses that affect human beings (cold, flu, herpes, papilloma etc.) naturally evolved towards sub-lethality to ensure their own diffusion within the population and avoid extinction: asymptomatic variants have a longer infection window and as a consequence higher spread compared with the lethal ones^13^. The evolution of the virus to sub-lethality is accelerated when the lethal variants of the virus are isolated, as happened after the admission of the hosts to the ICU. In this case, the isolation of the host, led to the suppression of the virus lethal variants.

In conclusion, our results highlight that predictive mathematical models, despite being useful to plan and implement effective countermeasures, cannot anticipate the human’s ability to identify and develop novel strategies to contain the spreading of the infection, nor the evolution of the virus. Human beings are not passive victims of the virus, but active fighters able to change the course of the infection using prevention and research to create adaptive strategies against the pandemic. To avoid the dramatic prediction of the SIDARTHE model, each country can, and should, take effective measurements to counteract the virus. We are not suggesting in any way that we can avoid to wear face masks or to keep social distancing, or to follow the rules. Rules should be strictly followed to avoid an increase of contagion. These data however underline that the Italian model, at present, is effectively working to constraint mortality and select the virus toward sub-lethal expressions. In other words, we can win.

## Data Availability

The Italian official data are available at the following link: https://github.com/pcm-dpc/COVID-19
The Matlab source code for this paper, including the modified algorithms from the original publication, can be found at the following link: https://drive.google.com/drive/folders/1EB1oLlxsWI7Xjs-5p6Zd-xrIz6fbkV1a?usp=sharing

## Authors Contributions

C.S., G.M., A.S.C.C. conceived the experiment; G.M. analyzed the data; C.S. drafted the manuscript; All the authors provided first-hand insight into the disease evolution and provided the clinical contextualization and interpretation of the results. All authors wrote and approved the manuscript.

## Additional Information

- For a detailed description of the methods please refer to Giordano et al. 2020^1^
- The Italian official data are available at the following link: https://github.com/pcm-dpc/COVID-19
- The Matlab source code for this paper, including the modified algorithms from the original publication, can be found at the following link: https://drive.google.com/drive/folders/1EB1oLlxsWI7Xjs-5p6Zd-xrIz6fbkV1a?usp=sharing

